# Deep Learning Model Using Continuous Skin Temperature Data Predicts Labor Onset

**DOI:** 10.1101/2024.02.25.24303344

**Authors:** Chinmai Basavaraj, Azure D. Grant, Shravan G. Aras, Elise N. Erickson

## Abstract

**Background:** Changes in body temperature anticipate labor onset in numerous mammals, yet this concept has not been explored in humans.

**Methods:** We evaluated patterns in continuous skin temperature data in 91 pregnant women using a wearable smart ring. Additionally, we collected daily steroid hormone samples leading up to labor in a subset of 28 pregnancies and analyzed relationships among hormones and body temperature trajectory. Finally, we developed a novel autoencoder long-short-term-memory (AE-LSTM) deep learning model to provide a daily estimation of days until labor onset.

**Results:** Features of temperature change leading up to labor were associated with urinary hormones and labor type. Spontaneous labors exhibited greater estriol to α-pregnanediol ratio, as well as lower body temperature and more stable circadian rhythms compared to pregnancies that did not undergo spontaneous labor. Skin temperature data from 54 pregnancies that underwent spontaneous labor between 34 and 42 weeks of gestation were included in training the AE-LSTM model, and an additional 40 pregnancies that underwent artificial induction of labor or Cesarean without labor were used for further testing. The model was trained only on aggregate 5-minute skin temperature data starting at a gestational age of 240 until labor onset. During cross-validation AE-LSTM average error (true – predicted) dropped below 2 days at 8 days before labor, independent of gestational age. Labor onset windows were calculated from the AE-LSTM output using a probabilistic distribution of model error. For these windows AE-LSTM correctly predicted labor start for 79% of the spontaneous labors within a 4.6-day window at 7 days before true labor, and 7.4-day window at 10 days before true labor.

**Conclusion:** Continuous skin temperature reflects progression toward labor and hormonal status during pregnancy. Deep learning using continuous temperature may provide clinically valuable tools for pregnancy care.

## Introduction

Body temperature reflects female mammalian reproductive status from adolescence^1^ through adult fertility^2–4^ and menopause.^3,5^ Body temperature monitoring is also of increasing interest in human pregnancy,^6^ as temperature change predicts parturition in a variety of species^7–10^ (**Table 1**). Historically, temperature measurements were most frequently taken from the “core”, requiring probes inside the body. However, *skin* surface temperature has emerged as a practical metric for monitoring the female reproductive system via its influence on thermoregulation and autonomic tone.^11–13^ Human skin temperature has already been deemed useful in cases ranging from peri-ovulatory window prediction^2,3,14^ to conception^15^ and fever detection.^16–20^

**Table 1:** Reported changes in body temperature (skin or core) among various mammalian species during pregnancy that have been observed prior to the onset of parturition.

| Species | Observed Change (T) | Time Window | Measurement and Frequency |
| --- | --- | --- | --- |
| lion<sup>35</sup> | -1.3 C | “late gestation” | intraperitoneal, ~ continuous |
| squirrel<sup>21</sup> | -1.2 C | -20 days | intraperitoneal, 1/min |
| orca whale<sup>36</sup> | -0.3 C, -0.8 C | -5 days, -24 hours | rectal, 1/day |
| wolverine<sup>37</sup> | -0.8 C | - 24 hours | intraperitoneal, 1 & 15 min |
| rabbit<sup>38</sup> | -0.7 C | < - 24 hours | intraperitoneal, 1 & 6 min |
| rat<sup>39,40</sup> | ~-0.5 C | -5 to -1 days | intraperitoneal |
| horse<sup>41</sup> | -0.5 c (-0.1) | (-24) -15 to -3 hours | rectal, 2/day |
| sheep<sup>42</sup> | -0.5 c | -24 hours | neck and vulvar, 1 & 10 min |
| cow<sup>8,9,43</sup> | -0.3- -0.2 C | -2.5 to 0 days | intravaginal; or ruminal |
| dog<sup>44</sup> | ≤ -0.3 C | -24 hours | intravaginal, daily means. |
| Moose<sup>45</sup> | ≤ -0.2 C | -3 to 0 days | ingested logger, 1 & 5 min |
| mouse<sup>46</sup> | <0.5 C | -72 h to -24 h | intraperitoneal, 1/min |
| goat<sup>10</sup> | not reported | n/a | vulva, 1/day |
| macaque<sup>47</sup> | not reported | -1 to -1.5 hours | subscapular, 1/min |

Both the central nervous system and peripheral vasculature contribute to the utility of temperature in reproductive monitoring. Briefly, estradiol promotes peripheral vasodilation, and the addition of progesterone leads to peripheral vasoconstriction.^11^ Estradiol therefore allows body heat to escape, lowering both core and skin temperature in females^21–23^ and progesterone, with or without the presence of estradiol, traps heat and increases metabolic rate, raising core and skin temperature.^3,4,14,24,25^ In addition, it was recently hypothesized that mechanisms linking thermoregulation and reproduction may originate centrally in the hypothalamus and ventral tegmental area.^12,22,23^ This phenomenon informs the basis for self-monitoring of the ovulatory cycle, with temperature tracing the trajectory of estrogen and progesterone production. Importantly, changes not only in temperature level but also in the cyclicity of temperature over hours and days (i.e., biological rhythms) indicate the peri-ovulatory period and pregnancy onset.^3,26–29^ Further, observed changes in temperature cyclicity mirror those in the levels and patterns of underlying reproductive hormones.^30^

While hormone patterns and sources (e.g., the placenta) differ in the third trimester of pregnancy from non-pregnant female neuroendocrine states, we hypothesize that the same principles apply to the role of steroid hormones on thermoregulation occurring prior to labor. Briefly, the influence of progesterone retreats in preparation for labor with rises in the ratio of estriol to progesterone,^31^ prolactin, corticotropin releasing hormone, and pro-inflammatory prostaglandins/ cytokines.^32–34^ Given the complexity of this hormonal state we might propose that changes in the ratio of progesterone and estriol would prompt decreasing temperatures, however, changes in the latter three factors might also suggest rising temperatures. Despite this lack of clarity as to what patterning pre-parturition hormones would produce, coarse decreases in body temperature have been noted with surprising consistency prior to labor in a wide range of mammalian species (**Table 1**).

Despite the reliability of these observations across phases of reproductive life and across species, the use of body temperature for predicting human labor onset has not been robustly studied. Presently, human parturition is estimated to occur within a range of weeks around a population mean of 40.0 weeks from the last menstrual period (an estimated 38 weeks post-conception), or via ultrasound performed in the first trimester which provides the gestational age of the embryo.^48^ However, present methods are associated with multiple weeks of average error^49^ based on natural variation in gestation length, reporting error, and variability in the timing of both ovulation and conception relative to the last menstrual period.^50^ Moreover, an individual’s length of gestation has not been predictable, despite attempts using AI/ML trained mostly on clinical features and ultrasound measures of cervical changes.^51–54^ It may be that a more appropriate method is the construction of a model that can detect subtle patterning in an output known to change before mammalian parturition: body temperature.

We previously demonstrated that multi-modal, daily markers can differentiate pregnancies destined to pass the clinical estimated ‘due date’ (EDD) from those laboring earlier.^55^ Our study, <u>Bio</u>logical rhythms <u>B</u>efore and <u>A</u>fter <u>Y</u>our <u>B</u>irth (BioBAYB); gathered daily temperature, activity, heart, and sleep data from wearable devices worn by participants in the third trimester prior to the EDD. A boosted random forest machine learning model was able to differentiate between pregnancies that would eventually pass the EDD versus those that would spontaneously deliver prior to the EDD. Although these single daily time points were not able to generate a precise due date estimate, we noted that trends in a sleeping average body temperature ranked consistently near the top of the features in the random forest across validation runs. The accuracy of this model was determined via the area under the receiver operating curve at 0.71, which denotes moderate ability to predict the outcome of longer versus shorter gestation. As the sample was comprised of mostly healthy and low-risk participants’ data, we were limited to term pregnancy predictions. Together, despite sampling limitations, the study indicated that a derived metric of temperature change was relevant to generating predictions when compared to heart rate, heart rate variability, sleep, and activity. This finding agreed with previous work^11,14,15,55^ suggesting that temperature is uniquely valuable in paralleling female reproductive status **(e.g., Figure 1)**, and that temperature metrics tuned specifically for the application of labor prediction may be more valuable still.

**Figure 1.**
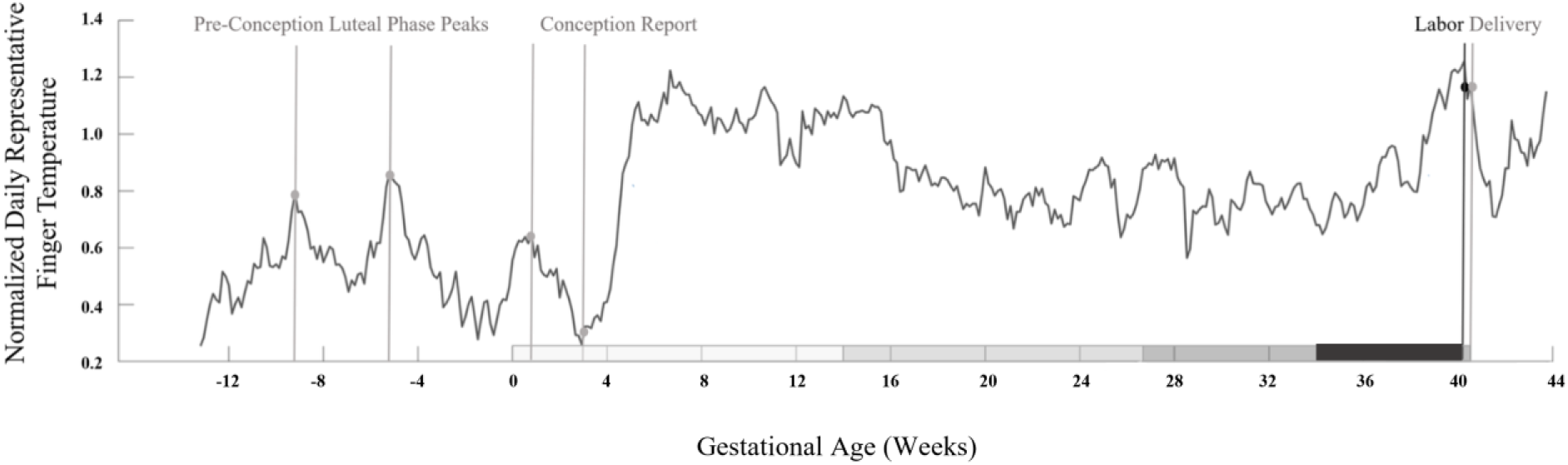
Normalized daily finger temperature sample from preconception to delivery. Normalized daily finger temperature sample from a representative spontaneously laboring mother. Black lines and dots indicate labor onset. Gray lines and dots indicate progression from 3 ovulatory cycles’ luteal phase temperature peaks, to reported conception, to eventual delivery. Gray bars on x-axis delineate trimesters, and the final black horizontal bar indicates the time period of data ultimately utilized in this dataset to build the labor-onset prediction model. A rise in skin temperature is observed beginning approximately 2-3 weeks prior to labor onset, with a reversal to a smaller drop in temperature within a week prior to labor onset. Reproduced with permission.^56^

In the present study, we examine the feasibility of using continuous body temperature for predicting human labor onset. We approach the problem both by relating hypothesis-driven changes in temperature to labor type and hormone levels, and by employing machine learning methods designed for rich time series. These data improve upon extant work by employing artificial intelligence methods specifically designed for time series, and in comparing observed features of temperature change preceding labor to change in hormonal trajectory using urinary hormone metabolites.

We hypothesize that labor onset predictions based on continuous skin temperature using advanced deep learning will yield greater accuracy than the current clinical EDD. We estimate that, comparable to other species, temperature will decrease as labor approaches. We also anticipate that the utility of temperature features will be reduced for participants with an induction or cesarean date (or an exogenous influence rather than physiological end to pregnancy). Finally, we hypothesize that features of body temperature’s change over time will be associated with changes in estrogen and progesterone urinary metabolites.

## Methods

### Experimental Design

#### Ethical Approval

The protocol for the original study was approved by an Institutional Review Board at Oregon Health & Science University and further via a Data Use Agreement with the University of Arizona for analyses of the de-identified dataset previously collected.

#### Sampling Method and Enrollment

Participants were recruited from maternity clinics as well as national social media advertising and enrolled following written informed consent. Inclusion was limited to adults who could provide written consent in English who were having a generally healthy pregnancy (no current hypertension or gestational diabetes and pre-pregnancy body mass index of less than 40 kg/m^2^) and anticipating a vaginal birth. Those already planning to undergo labor induction at less than 41 weeks, who had ovulatory dysfunction, uncontrolled thyroid disorder, or who used in-vitro fertilization, as well as those working night or rotating shifts were excluded. A second cohort of participants was recruited with same criteria as above though also had risk factors for preterm birth (e.g., multiple gestation, history of spontaneous preterm birth).

#### Study Procedures

Participants were fitted to their ring size using a ring-fitting kit provided by the ring manufacturer (Ouraring Inc.) and were instructed to wear the ring as continuously as possible throughout the remainder of the pregnancy, on whichever finger achieved the best fit on the non-dominant hand. REDCap surveys were used to gather self-reported pregnancy symptoms, clinical assessment data, labor and birth events, and psychometric tools as previously reported.^55^

#### Temperature Data Collection

The Ouraring is a commercial health tracking device worn on the finger. The Gen2 Ouraring is equipped with temperature (negative temperature coefficient (NTC) derived from 3 thermistors), 3-D accelerometer, and infrared photoplethysmography (PPG) sensors and measures physiological signals, such as heart rate (HR), heart rate variability (HRV), per minute finger temperature, respiration, and movement. The sensors are housed in the inner part of the ring on the palm side of the finger. Data is transmitted from the ring to the user’s phone via Bluetooth, and from the phone it is uploaded to the cloud. Continuous data collection enabled the establishment of personalized biometric baselines for each user. Continuous finger temperature data, collected over a branch of the brachial artery, is the exclusive subject of the present study. Results based on other outputs collected by the device have been previously reported.^55^

#### Data Acquisition Pipeline

Following the report of the participant’s delivery, data was downloaded from the cloud into secure cloud storage through the research institution. **Supplemental Figure 1** outlines our data ingestion and storage architecture. Data was made available through SQL queries and was accessed through the SensorFabric Python library (created by University of Arizona Sensor Analysis Core^57^).

### Raw temperature and hormone analyses

#### Participant Self-Collection of Urine Samples

A subset of 30 participants self-collected a first morning urine sample in a plastic basin at home each morning beginning at 38 weeks of gestation. Dried Urine Test for Comprehensive Hormones® (DUTCH) (Precision Analytical Inc., McMinnville, OR) test strips (2×3 in sized Whatman body fluid collection paper) were dipped into the urine and allowed to dry completely for 24 hours before storage in a collection bag in a home freezer. Participants aimed to sample every day from 38 weeks until onset of labor. We analyzed the up to 10 samples (as available) prior to labor onset per participant. Each specimen was assayed for the following: Estrone (E1), Estradiol (E2), Estriol (E3), α- and β-pregnanediol (αPg and βPg; the main progesterone metabolites found in urine), cortisol, and melatonin.

#### Hormone Assay

As previously reported,^3,58^ estrogens, αPg and βPg, cortisol and melatonin were analyzed using DUTCH test’s proprietary in-house assays on the Agilent 7890/7000B gas chromatography-mass spectrometry (GC–MS/MS) (Agilent Technologies, Santa Clara, CA, USA). The equivalent of approximately 600 μl of urine was extracted from the filter paper using acetate buffer and hydrolyzed to free forms with a reported >90% recovery. Creatinine was measured in duplicate using a conventional colorimetric (Jaffe) assay. Conjugated hormones were extracted (C18 solid phase extraction), hydrolyzed by Helix pomatia and derivatized prior to injection (GC–MS/MS) and analysis. The mean inter-assay coefficients of variation were 7.4% for E2, 14.9% for αPg, and 13.6% for βPg. The mean intra-assay coefficients of variation were 7% for E2, 12% for αPg and 12% for βPg. Sensitivities of the assays used were as follows: E2 and αPg, 0.2 ng/mL; βPg, 10 ng/ mL. Samples were examined with respect to a standard curve for expected range of concentrations and controls, and results were further normalized to creatinine in the samples.

#### Biological Rhythm Analysis

Potential changes to circadian power of skin temperature (mean power per minute within the 23–25 h band) were assessed. Wavelet Transform code was modified from the MATLAB Jlab toolbox and from Dr. Tanya Leise^59^ in MATLAB 2022b. In contrast to Fourier transforms that transform a signal into frequency space without temporal position (i.e., using sine wave components with infinite length), wavelets are constructed with amplitude diminishing to 0 in both directions from center. This property permits frequency strength calculation at a given position. Wavelets can assume many functions (e.g., Mexican hat, square wave, Morse); the present analyses use a Morse wavelet with a low number of oscillations (defined by *β* and *γ*), as in previous studies.^59,60^ Morse Wavelet parameters of *β* = 5 and *γ* = 3 describe the frequencies of the two waves superimposed to create the wavelet, as in previous studies.^25,61^ This low number of oscillations enhances detection of contrast and transitions.

#### Hormone Data Analysis and Temperature Comparisons

Hormone data was analyzed over the last 10 days leading up to labor. Datasets with only one data point were removed (n=2). Data were linearly interpolated, and normalized using the MATLAB function “normalize”, which ignored NAN values. Hormonal data were then compared to temperature metrics by individual and in aggregate. For the purposes of gross temperature and hormone data comparison, participants were labeled as trending up or trending down. Trending up was defined as someone whose 72-hour smoothed temperature time series sloped up over the last 10 days of pregnancy. Trending down indicates that smoothed temperature sloped down over the last 10 days of pregnancy. Temperature time series from participants who did urine sampling and experienced spontaneous labor (n=18) were divided into increasers and decreasers, and hormone time series were plotted between the two.

#### Standard Statistical Methods Comparing between Spontaneous and Induced/Pr-elabor Cesarean

Sample demographic and clinical characteristics were compared between the group experiencing a spontaneous labor to those undergoing labor induction or a prelabor Cesarean birth. We used bivariate parametric and non-parametric tests as indicated. For hormone and temperature time series features, values are reported as mean ± 95% confidence interval (C.I.). For statistical comparisons of temperature features, Friedman’s tests (non-parametric repeated measures ANOVAs) were used to assess differences between time series of spontaneous and induced labors. Kruskal Wallis (KW, nonparametric ANOVA) tests were used for comparisons of individual means by group, as indicated. Trends over time were assessed using Mann-Kendall tests. For Friedman’s and KW tests, χ2 and p values are listed in the text. Figures were formatted in Microsoft PowerPoint 2023 (Microsoft Inc., Redmond, WA) and Adobe Photoshop CS8 (Adobe Inc, San Jose, CA).

### Labor Prediction Methods

#### Participant inclusion criteria

Figure 2 details an overview of participant breakdown for model development. Of the total 127 participants enrolled 7 were lost to follow-up. Of the remaining 120 participants, 71 gave birth spontaneously, 45 had to be induced and 4 underwent Cesarean without labor onset. The model was trained on the spontaneous (including spontaneous labor with augmentation) group for which we knew the actual date and time of labor onset, unlike the non-spontaneous group. From the spontaneous group 17 participants were dropped for one of the following reasons (a) less than 21 days of contiguous skin temperature data prior to labor, (b) less than 75% data density, which is defined as a ratio that indicates the total missing skin temperature data in days due to non-wear and dead battery. Following the same criteria for non-spontaneous (except replacing labor day with induction or Cesarean day), 12 participants were eliminated from that group, including 1 Cesarean.

**Figure 2.**
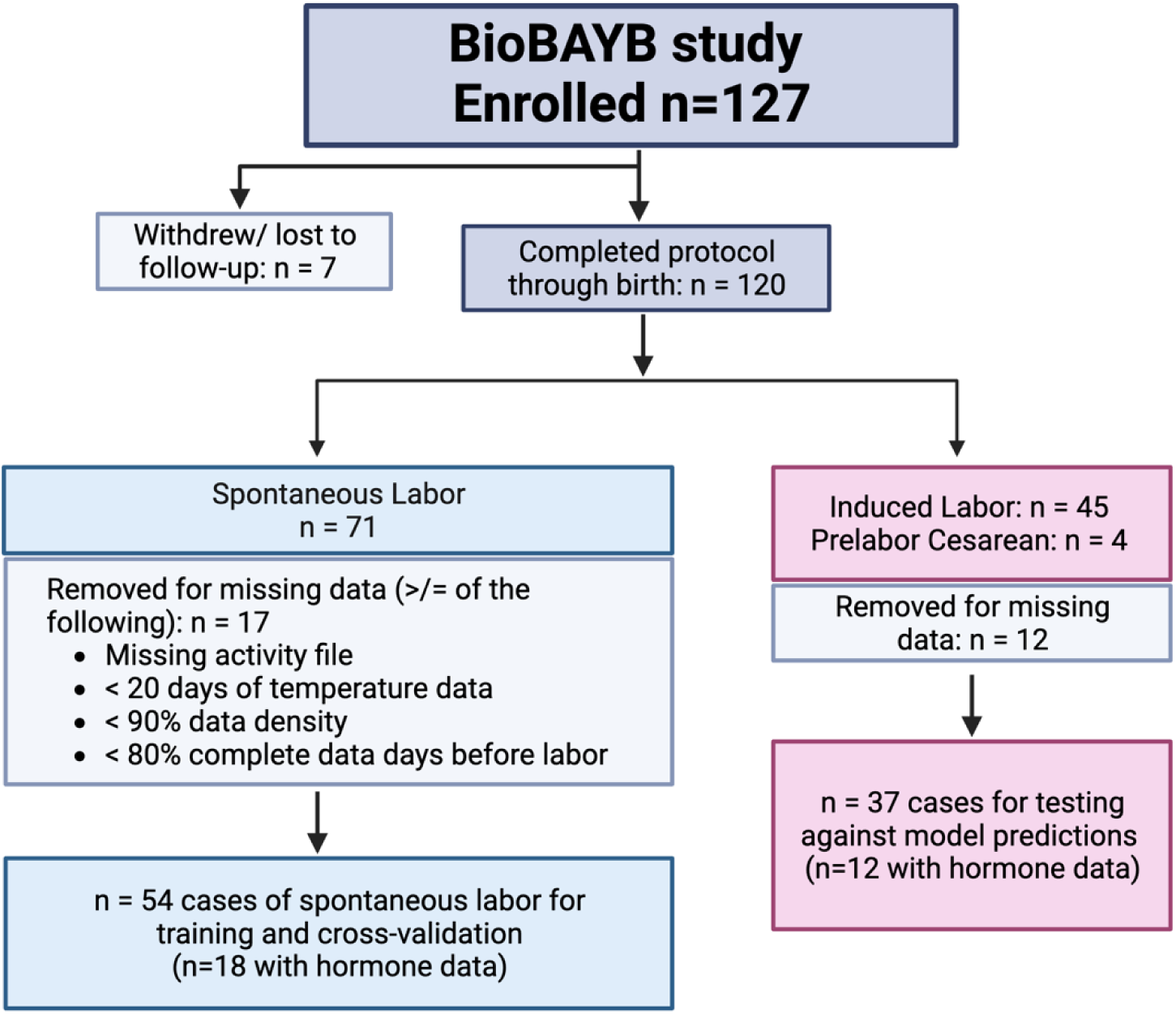
Diagram of Biological Rhythms Before and After Your Birth (BioBAYB) study participants through data cleaning stages and final analytic groups for training and cross-validation of the Auto Encoder Long-Short Term Memory model.

#### Data Preparation for Deep Learning

For all the participants, per-minute temperature data were averaged over a 5-minute window and segmented into 24-hour periods starting at 10 am each day, (**Supplemental Figure 1**). Next, sections of temperature data associated with non-wear, i.e. data collected when the user removed the ring were isolated and removed using the labelled 5-minute activity data obtained from the ring. Next, we used linear interpolation method to account for the missing and non-wear temperature data segments. Linear interpolation performed better than other interpolation methods such as – polynomial, cubic-spline, and inverse distance weighting in accounting for missing data.

#### AI Model Problem Definition

Our model was formalized as a non-linear approximator *F*: *Y*_*k*_ = *F*(*T*_*k*_, θ) + δ. Given the input sequence of daily temperatures, *T*_*k*_ = {*T*^*k*−1^, *T*^*k*−2^, *T*^*k*−3^ … *T*^1^} starting from the *k*^*th*^ day of gestation where *T*^*k*^ is a sequence of aggregate 5-minute temperatures for that day, our model is represented by a set of trainable parameters θ, and predicts a value *Y*_*k*_ that indicates number of days until labor relative to current gestational age *k*. δ represents the parameters of regularization employed to avoid model over-fitting. We train the model using an autoregressive approach, commonly used in time-series forecasting problems, where we predict days until labor (at *k*^*th*^day of gestation) using only skin temperature data from the past. We find model *F* over the space of all non-linear models that minimizes the Mean Absolute Error (MAE) objective function 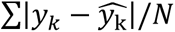 for all training subjects *N*, where 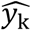 is the predicted days until labor and *y*_*k*_ is true days until labor at a gestational age of *k*. As deep neural networks (DNN’s) are highly effective universal non-linear approximators, we chose to use a combination of convolutional autoencoders (AE) coupled with a long short-term memory (LSTM) network to train model parameters θ and δ.

#### Auto Encoder Architecture

Autoencoders are excellent at converting input data from the feature space to latent space,^62^ reducing data dimensionality and in the process automatically reducing noise^63^ in the data. For our analysis, we developed a convolutional AE which uses convolutional layers as the basic building blocks. The AE is a special type of DNN which is trained to reconstruct its input data from a compressed latent representation. For example, in our case it first encodes the daily skin temperature data to an encoded latent representation, then decodes the latent representation back to daily skin temperature data while minimizing the reconstruction error. The encoder part of the AE generated a 64-dimensional latent space representation of the daily temperature data which was then used as the input for the LSTM. The daily temperature data from the same participant were treated as independent data points and the AE was trained on a total of 3000 daily temperature data points. The detailed architecture of the convolutional AE is summarized in **Supplemental Figure 2**. The AE was able to reconstruct the daily temperature data from the encoded representation with a mean error of 1.48℃.

#### Model Cross-validation and Evaluation

We used a subject-oriented approach to evaluate our model, where a participant’s data is wholly used either for training or testing. To evaluate the performance of all our models and methods, we employ a k-fold cross validation method, where we split the participants into *k* folds. We train the model using *k-1* folds of data and test the model performance on the held-out *k^th^*fold. We used k=9 which gave us 6 participants per fold. This process is repeated k times until each fold has served as the test dataset at least once. Subject-wise cross validation is a more effective way of evaluating model performance as it ensures that the model does not see any part of the test participants’ physiological data during training, and it also improves generalization to new participants’ data. As we are dealing with time series data, the performance metrics are evaluated as a function of time. We compute the mean difference and absolute mean difference of model prediction to the ground truth at a specific point in time and report the errors across time. We assess the AE-LSTM model performance by evaluating the mean absolute error in predicting labor onset relative to current gestational age with the true labor onset. We established valid prediction baselines based on the current standard using EDD and compared the AE-LSTM performance to the baseline models. To better interpret the model’s performance clinically, we considered all model predictions that go past the actual date of labor as negative (i.e., false-negative; the mother delivered earlier than predicted), and all predictions that were earlier than the date of labor as positive (i.e., false-positive; the mother delivered later than predicted) **(See Figure 6**, **Table 3**).

#### Converting prediction out from AE-LSTM to labor window

To improve clinical interpretation, we converted the predicted value returned by AE-LSTM (*Ŷ*_*T*_) at time *T* (days until labor) into window [*W*_1_, *W*_2_], such that model prediction falls within it (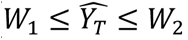) with a given probability *P*. This was achieved by converting the discrete prediction errors (MAE) for all participants during cross-validation across all folds at each time point *T* into a distribution *E*(*T*) by calculating a Kernel Density Estimate. We tested *E*(*T*) for normality using the Shapiro-Wilk test. With *E*(*T*) modeling a normal distribution with mean *Ŷ*_*T*_, the area under the KDE curve between bounds (ɛ_1_, ɛ_2_) then gave us the probability *P* of model error lying between them. We calculated the window bounds as *W*_1_ = *Ŷ*_*T*_ − ɛ_1_, *W*_2_ = *Ŷ*_*T*_ + ɛ_2_for a given probability *P* ∈ {0.7, 0.8, 0.9, 0.95} and reported the window size |*W*| = |*W*_1_| + |*W*_2_| across *Ŷ*_*T*_ within which the prediction was likely to fall. We define this window as W(P)_T_.

#### Prediction of future labor onset for participants with induced labors or pre-labor Cesarean birth

Labor induction may occur for many reasons and involves use of medications or mechanical devices to stimulate uterine contractions and cervical dilation. We chose to evaluate this non-spontaneous (exogenous) form of labor onset (or lack of labor for cases of pre-labor Cesarean) with the hypothesis that accuracy of predictions would be lower in this cohort. A model that performed with equal accuracy in pregnancies that entered labor naturally, compared to those that had an induction or Cesarean, would not likely be identifying features relevant for the physiological onset of labor. Separately, in this framework, induced pregnancies were interpreted as, “less ready for labor,” even at a late gestational age in comparison to individuals undergoing spontaneous labor. Therefore, we evaluated whether or not our predictions tended to fall later than actual induction within the induced cohort. Second, we evaluated whether induced labors’ prediction errors exhibited higher variability, as on average the physiological trajectory of the induced or complicated pregnancy should be different than the spontaneously laboring, healthy pregnancy.

## Results

### Sample Description

A total of 91 individuals were included. Spontaneous onset of labor occurred for 54 (59.3%) participants at a mean (standard deviation) gestational age of 39.9 (1.1) weeks and a range of 37.4 – 41.9 weeks, compared to mean of 39.5 (1.2) and a range of 36.1-41.2 among non-spontaneous labors/births. Mean gestational age did not differ between groups, nor did maternal age, EDD assignment method (ultrasound versus last menstrual period date), educational level, self-reported race or ethnicity, or sex of the infant. Seven participants (7.7%) reported developing gestational hypertension after enrolling in the study. Only one preterm birth was reported among labor induction group for an obstetric complication. Seven labor inductions were performed for pre-labor spontaneous rupture of membranes without a clear indication of labor starting at the time of induction, thus these individuals were categorized as labor induction and data was not used for training the model. For the other 33 labor inductions, 7 (21.2%) were attributed a post-dates pregnancy (i.e., passing the due date), 5 (15.2%) due to a medical complication, 10 (30.3%) indicated there was a fetal health indication and 4 (12.1%) chose a labor induction for convenience or other non-medical reason. **(Table 2).**

**Table 2.** Demographic and clinical features of BioBAYB participants by onset of labor (N=91).

|  | **Labor Group** |  |  |
| --- | --- | --- | --- |
|  | Total | Planned Cesarean or Induced Labor | Spontaneous Labor |
| **N** | 91 (100.0%) | 37 (40.7%) | 54 (59.3%) |
| **Maternal age, mean (SD) years** | 32.3 (3.5) | 32.2 (3.3) | 32.4 (3.6) |
| **Gestation at enrollment, mean (SD) weeks** | 30.2 (2.8) | 29.9 (3.0) | 30.4 (2.7) |
| **Gestation at birth, mean (SD) weeks** | 39.7 (1.1) | 39.5 (1.2) | 39.9 (1.1) |
| **Weight of newborn, mean (SD) lbs** | 7.7 (1.0) | 7.4 (1.0) | 7.9 (1.0)* |
| **Pre-pregnancy BMI, mean (SD) kg/m<sup>2</sup>** | 23.8 (4.0) | 24.7 (4.6) | 23.1 (3.4) |
| **Method for determining EDD** |  |  |  |
| Last menstrual period | 69 (76.7) | 27 (75.0) | 42 (77.8) |
| Ultrasound before 12 weeks | 16 (17.8) | 8 (22.2) | 8 (14.8) |
| Ultrasound after 12 weeks | 4 (4.4) | 1 (2.8) | 3 (5.6) |
| Conception date | 1 (1.1) |  | 1 (1.9) |
| **Parity** |  |  |  |
| Nulliparous | 48 (52.7%) | 25 (67.6%) | 23 (42.6%)* |
| Multiparous | 43 (47.3%) | 12 (32.4%) | 31 (57.4%) |
| **Educational Level** |  |  |  |
| High school graduate | 2 (2.2%) | 0 (0.0%) | 2 (3.7%) |
| Some college | 3 (3.3%) | 3 (8.1%) | 0 (0.0%) |
| College graduate | 43 (47.3%) | 15 (40.5%) | 28 (51.9%) |
| Graduate/Masters | 32 (35.2%) | 15 (40.5%) | 17 (31.5%) |
| Doctorate/professional degree | 11 (12.1%) | 4 (10.8%) | 7 (13.0%) |
| **Self-reported race/ethnicity** |  |  |  |
| White/European, non-Hispanic | 77 (85.6%) | 31 (86.1%) | 46 (85.2%) |
| Black / African, non-Hispanic | 1 (1.1%) | 0 (0.0%) | 1 (1.9%) |
| Hispanic | 5 (5.6%) | 2 (5.6%) | 3 (5.6%) |
| Asian | 7 (7.8%) | 3 (8.3%) | 4 (7.4%) |
| **Sex of infant (at enrollment)** |  |  |  |
| Male | 37 (40.7%) | 19 (51.4%) | 18 (33.3%) |
| Female | 36 (39.6%) | 14 (37.8%) | 22 (40.7%) |
| Unknown | 18 (19.8%) | 4 (10.8%) | 14 (25.9%) |
| **Location of birth** |  |  |  |
| Hospital | 79 (86.8%) | 37 (100.0%) | 42 (77.8%)** |
| Home | 7 (7.7%) | 0 (0.0%) | 7 (13.0%) |
| Birth center, non-hospital | 5 (5.5%) | 0 (0.0%) | 5 (9.3%) |
| **Labor Progress** |  |  |  |
| Spontaneous onset and progress | 34 (37.4%) | 0 (0.0%) | 34 (63.0%*** |
| Spontaneous onset/ augmented | 20 (22.0%) | 0 (0.0%) | 20 (37.0%) |
| Labor induction | 34 (37.4%) | 34 (91.9%) | 0 (0.0%) |
| Cesarean birth pre-labor | 3 (3.3%) | 3 (8.1%) | 0 (0.0%) |
| **Mode of birth** |  |  |  |
| Vaginally | 72 (79.1%) | 27 (73.0%) | 45 (83.3%)* |
| Vaginally with vacuum or forceps | 4 (4.4%) | 0 (0.0%) | 4 (7.4%) |
| Cesarean | 15 (16.5%) | 10 (27.0%) | 5 (9.3%) |
| **Postpartum hemorrhage** | 7 (7.7%) | 3 (8.1%) | 4 (7.4%) |
| **Hypertension during pregnancy** | 7 (7.7%) | 4 (10.8%) | 3 (5.6%) |

\*= $p < 0.05$ , \*\* $p < 0.01$ , \*\*\* $p < 0.001$ , BMI, Body Mass Index; EDD, Estimated Date of Delivery.

### Hormone metabolites link the mechanisms of labor onset to skin temperature

Spontaneous labors exhibited decreasing temperature (**Figure 4A**), circadian power (**Figure 4B),** and normalized α-Pregnanediol (**Figure 4C**) in the week prior to labor onset (median Mann-Kendall p=7.41*10^-8^, p=5.57*10^-9^, p=0.028; respectively). Although the typical spontaneous trend was decreasing temperature prior to labor, not every individual exhibited this trend (data not shown). α-Pregnanediol concentration in the week prior to labor onset differed by whether or not individuals exhibited downward sloping (grey, n=10) or showed no trend/upward sloping (black, n=8) temperature across that window, with those with in the falling temperature group trending toward lower median α-Pregnanediol (**Figure 4D)** (p=0.045). Temperature levels and hormone concentrations revealed additional differences between spontaneous and induced labors. Spontaneous labors exhibited colder temperatures by an average of 0.4 °C (**Figure 5A)** and greater circadian power (**Figure 5B)** (Friedman’s χ^2^=346, 240; p=1.7*10^-59^, 4.12*10^-38^, respectively). Finally, spontaneous labors exhibited a greater mean ratio of estriol to α-Pregnanediol across this window (a previously hypothesized marker of successful labor^64^) (KW χ^2^=14.3, p=2*10^-4^) (**Figure 5C**). Taken together with the downward trend of temperature and pregnanediol in anticipation of labor, these data support that spontaneous labors studied here may be in a later “physiological” stage of pregnancy and that this state is observable via body temperature. Together, thermoregulation and hormonal state appear to undergo related changes in preparation for labor.

**Figure 3.**
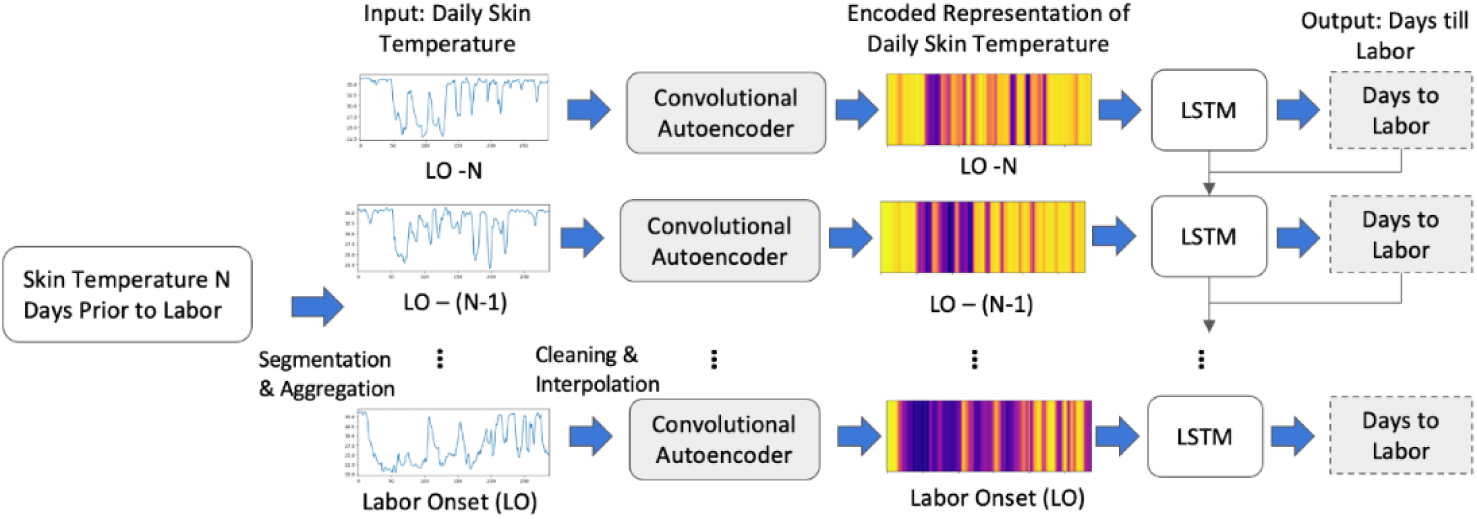
AE-LSTM Model Architecture Extracts Temperature Features Relevant to Labor. Contiguous daily skin temperature data is fed into a convolutional autoencoder, which outputs an encoded representation of length 64 for each day. Heatmaps depict the actual encoded representation of sample days of data. These are then fed into an LSTM (Long-Short Term Memory) in an autoregressive fashion to obtain a “days until labor onset” values relative to the current gestational age.

**Figure 4.**
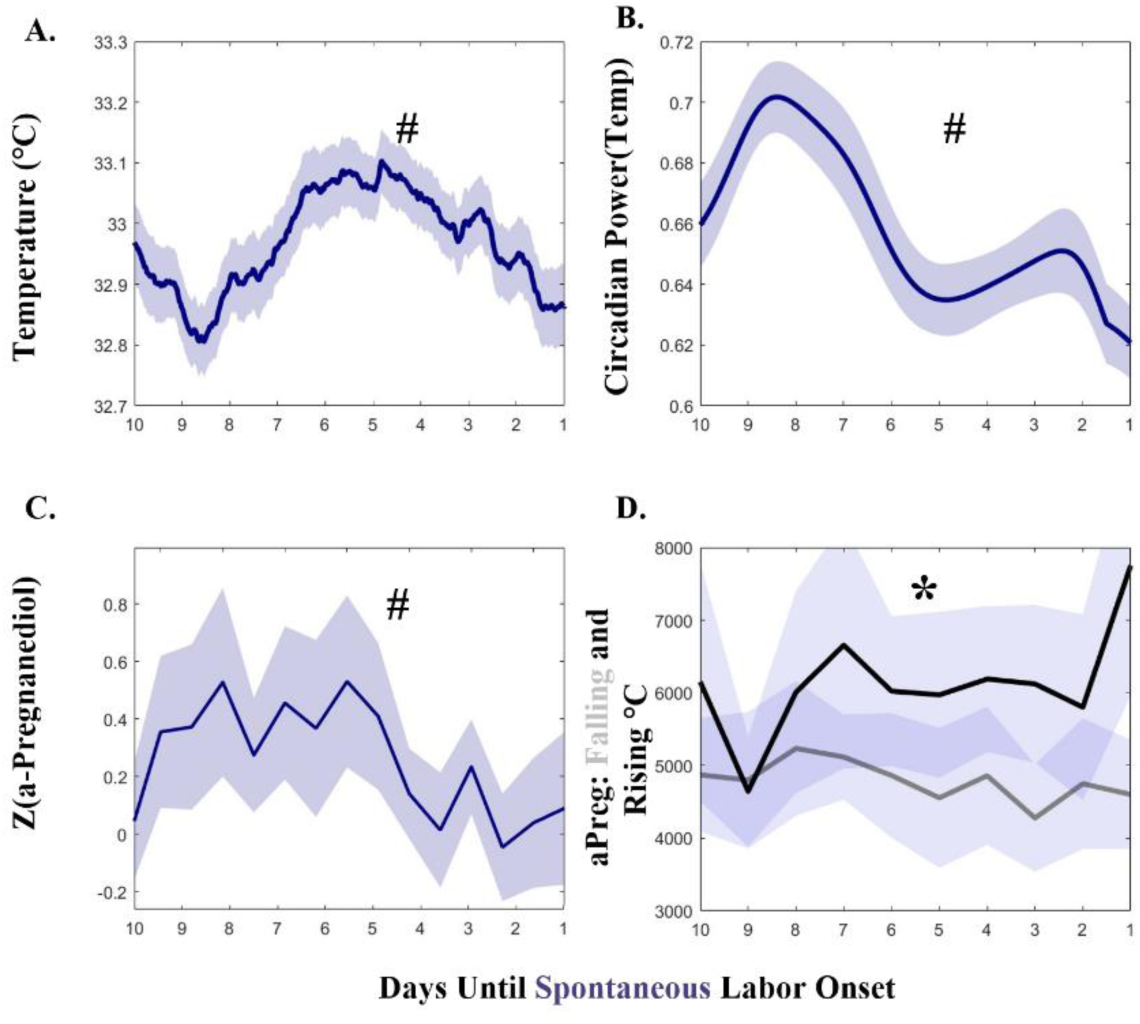
Temperature, Temperature Circadian Power, and αPregnanediol Exhibit Parallel Decreases Approaching Labor. Temperature level (A) and circadian power (C) parallel pattern of α-Pregnanediol (B) in the 10 days prior to labor onset. (D) α-Pregnanediol is reduced in individuals that exhibit falling temperatures in the 10 days prior to labor onset (D, gray line), as opposed to those who exhibit rising temperatures across that window (D, black line). Symbols: # indicates statistical Mann-Kendall trend over time in the week prior to labor; * indicates statistical difference between α-Pregnanediol in pregnancies with and without decreasing temperature in the week prior to labor. All solid lines are means and all error bars represent ± 95% C.I.

**Figure 5.**
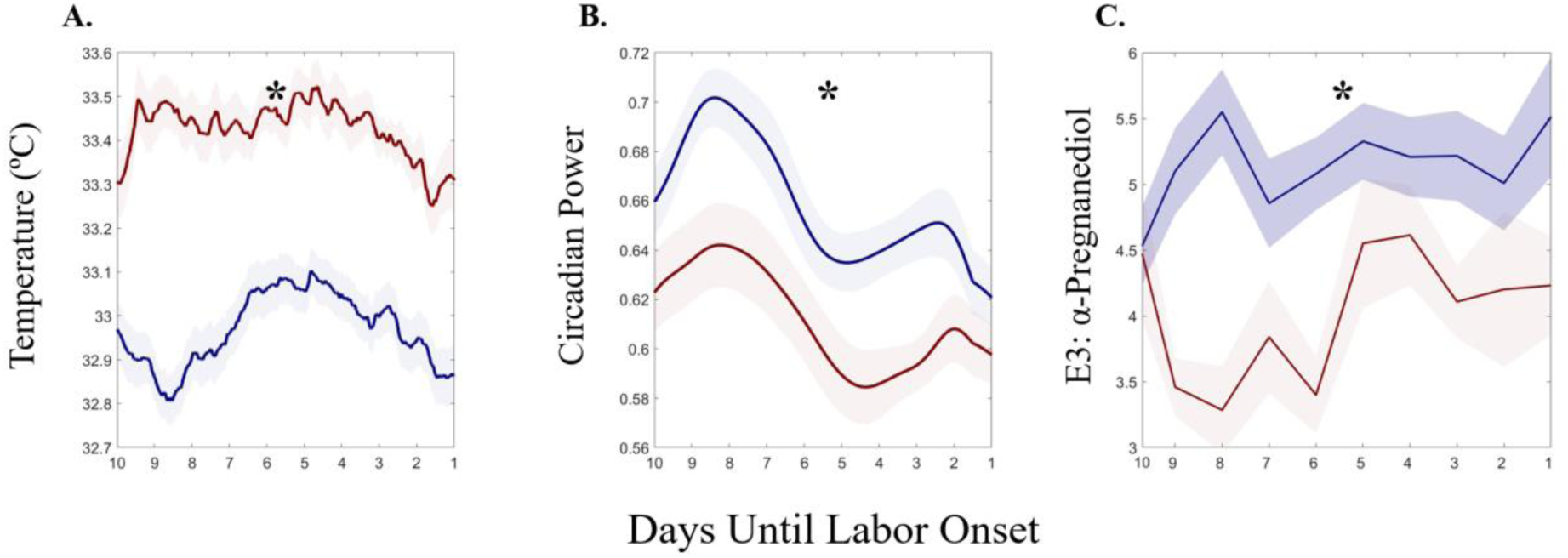
Spontaneous Labors Exhibit Lower Temperatures, More Stable Circadian Rhythms, and Greater E3:αPregnanediol Ratio. **A).** Body temperature in spontaneous (blue) and induced (red) labors. B). Wavelet circadian power is greater in spontaneous (blue) pregnancies. D) Spontaneous labors exhibit a greater ratio of E3: α-Pregnanediol. All solid lines are means and all error bars represent ± 95% C.I.

### AE-LSTM Model Accurately Anticipates Spontaneous Labor Onset

Model signed mean error decreased in participants who went on to spontaneously start labor from 40 to 25 days to labor onset after which it varied between 0 to 2 days (**Figure 6A**, **blue**). This error rate was not dependent on gestational age in spontaneous labors (n.s. Mann Kendall) (**Figure 6B, blue** and was lower than the clinical due date error of 12 days for the population at all time points. Average signed error rate was greater in pregnancies with induced labors (**Figure 6A and B, red)**, and trended downward with advancing days toward induction as well as with advancing gestational age (Mann Kendall p<0.001). Average signed error reached its minimum value and minimum variability for spontaneous labors at 1 week prior to labor onset with a mean (SD) of -0.3 (2.1), as compared to -2.8 (6.7) days for inductions. Error skewed 2 days late in spontaneous labors, on average, versus 10 days late for labor inductions across the entire prediction window. Mean signed prediction error by individual ranged widely between the groups, with induced labor ranging over 25 days of error and spontaneous labor ranging over 10 days.

**Figure 6.**
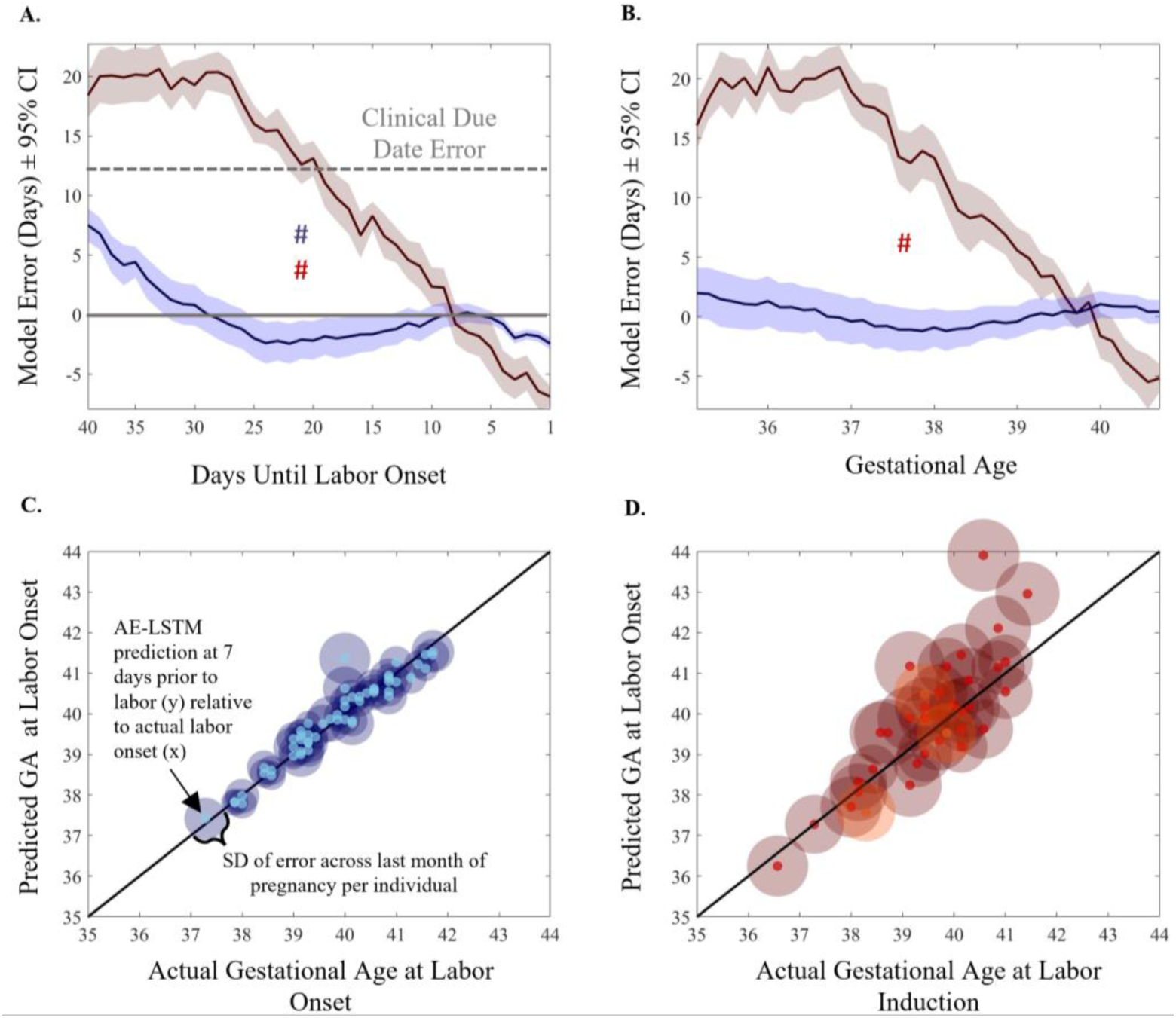
AE-LSTM Predicts Spontaneous Labor Onset With < 2 Days of Error in the Last 8 Days of Pregnancy. **Independent of Gestational Age.** (A). Spontaneous model error in days, ± 95% C.I. for the population for predictions generated each day. # Indicates statistical Mann-Kendall trend over time. Error decreases statistically across the window from labor -40 days to the day before labor onset. Gray dashed line indicates population mean error in days for the traditional due date based on last menstrual period. Gray solid line marks 0 days error. (B). Spontaneous model signed error in days. ± 95% C.I. for predictions generated at each gestational age indicates that spontaneous model performance (blue) does not vary statistically by gestational age at prediction. (C). Spontaneous labor error is lower and less variable than induced error across gestational ages, compared to Induced error (D). Spontaneous data are plotted in blue; induced data are plotted in red. Three available Cesarean births are shown in orange. Transparent circle diameter is proportional to that individual’s signed model error across the month prior to labor onset, whereas the solid center circle represents the minimum model error for that individual.

**Figure 7.**
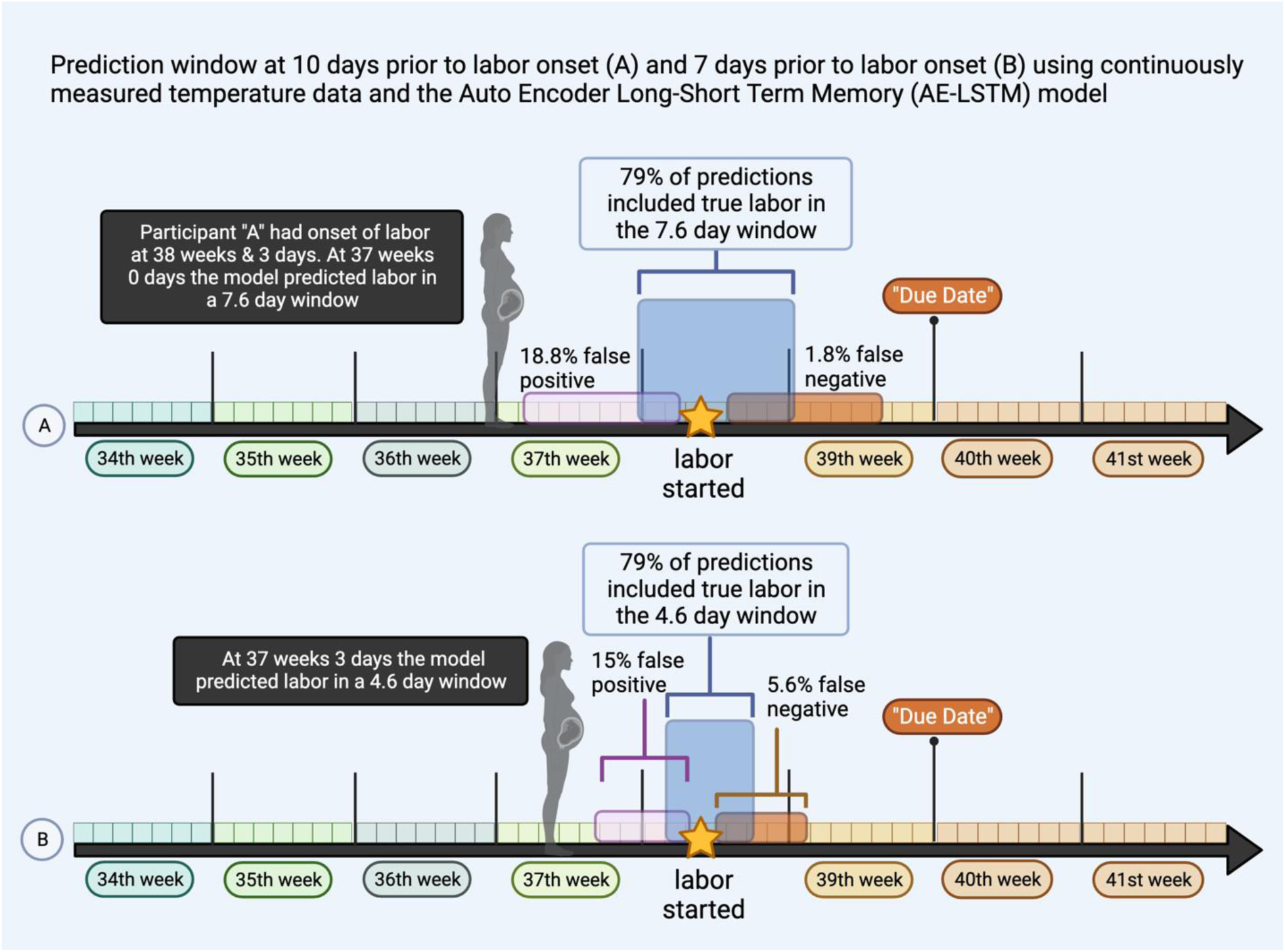
Graphical Summary of AE-LSTM model with labor predictions in a future window of time relative to true labor onset. A) at 10 days prior to labor onset, the model predicts a window of 7.6 days that accurately included true labor date in 79% of the sample. The window was before true labor 18.8% of the time with a mean (SD) of 0.6 (0.5) days away from labor onset. Conversely the predictions resulted in a false negative in 1.8% of cases (n=1) in that labor occurred prior to the predicted window to occur 0.9 days after labor actually started. B) Illustrates that at 7 days prior to labor, the window shrinks to a smaller range of days with similar rates of accurate prediction.

Scatters of actual gestational age at delivery versus predicted gestational age at delivery at 1 week prior to labor onset for all participants illustrate increased prediction accuracy in individuals who had spontaneous labor (linear model fit R^2^=0.93, AIC = -134) (**6C, light blue dots**). By contrast, scatter of actual gestational age at induction relative to predictions made 1 week prior to induction reveal reduced accuracy (R^2^=0.68, AIC=-8.81) (**6D, bright red dots**). Moreover, data illustrate lower individual SD of prediction accuracy over the last month of pregnancy (**6C, dark blue shaded region diameter)**, as compared to the larger SD of induced predictions (**6D, dark red shaded region diameter**). Data for the 3 planned Cesarean births are visualized (**6D, orange dots**), with error and error variability comparable to inductions (error at 1 week prior to labor of -7.3, 4.9, and 2.3 days, respectively; SD= 6.5 days, error range = 36 days over month prior to delivery). Together, these findings support that induced labors may be less physiologically ready for labor, that it is more difficult to generate consistent predictions about their pregnancy progression, and that in the absence of labor readiness, advancing gestational age (as measured via advancement across the temperature time series) may be a salient predictor of approximately when labor should occur.

### Distribution of Error Across Cross-Validation Folds at 7 Days

We choose 7 days before as a representative cross-section to showcase model error across various folds during cross-validation. Table 3 shows an overview distribution of model error across all folds for 7-day cross-section, with each fold containing 6 participants that underwent spontaneous labor. The overall error across all folds at 7 days was μ=-0.08, α=1.63 days. We observed an overall closely clustered error distribution across all folds. Folds 1 showed the largest mean error of -1.93 days followed by fold 2. While fold 5 showed the smallest mean error of -0.17, it also showed a wider spread of error distribution α=2 days greater than any other fold.

**Table 3.** Distribution of error for each fold at 7 days prior to true labor during model cross-validation.

| Folds | 1 | 2 | 3 | 4 | 5 | 6 | 7 | 8 | 9 |
| --- | --- | --- | --- | --- | --- | --- | --- | --- | --- |
| Errors | -1.93 | -1.27 | -0.92 | -0.73 | -0.17 | 0.3 | -0.34 | 0.86 | 0.65 |
| (days) | $\pm 1.8$ | $\pm 1.1$ | $\pm 1.14$ | $\pm 0.7$ | $\pm 2$ | $\pm 1.6$ | $\pm 1.25$ | $\pm 1.22$ | $\pm 1.32$ |

**Table 4.**
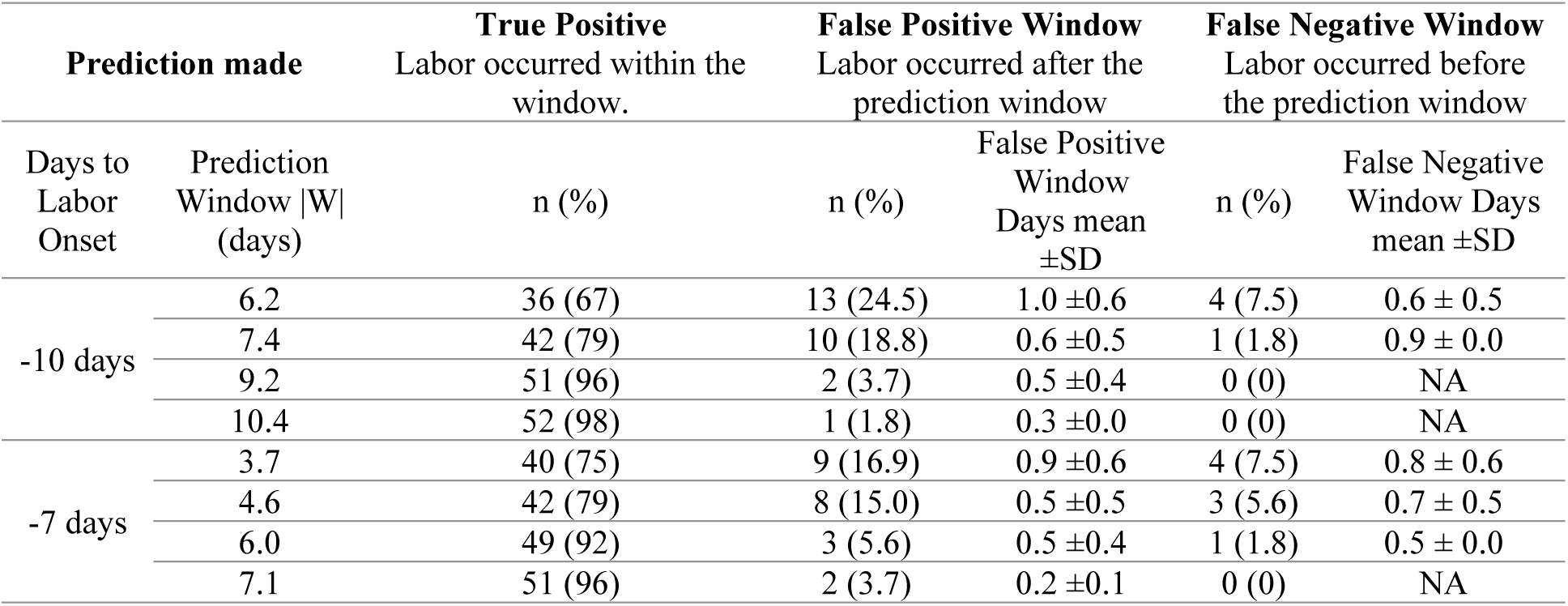
Model Prediction Accuracy Windows in Spontaneous Labor.

| Prediction made |  | True Positive<br>Labor occurred within the<br>window. | False Positive Window<br>Labor occurred after the<br>prediction window |  | False Negative Window<br>Labor occurred before<br>the prediction window |  |
| --- | --- | --- | --- | --- | --- | --- |
| Days to<br>Labor<br>Onset | Prediction<br>Window \|W\|<br>(days) | n (%) | n (%) | False Positive<br>Window<br>Days mean<br>±SD | n (%) | False Negative<br>Window Days<br>mean ±SD |
| -10 days | 6.2 | 36 (67) | 13 (24.5) | 1.0 ±0.6 | 4 (7.5) | 0.6 ± 0.5 |
|  | 7.4 | 42 (79) | 10 (18.8) | 0.6 ±0.5 | 1 (1.8) | 0.9 ± 0.0 |
|  | 9.2 | 51 (96) | 2 (3.7) | 0.5 ±0.4 | 0 (0) | NA |
|  | 10.4 | 52 (98) | 1 (1.8) | 0.3 ±0.0 | 0 (0) | NA |
| -7 days | 3.7 | 40 (75) | 9 (16.9) | 0.9 ±0.6 | 4 (7.5) | 0.8 ± 0.6 |
|  | 4.6 | 42 (79) | 8 (15.0) | 0.5 ±0.5 | 3 (5.6) | 0.7 ± 0.5 |
|  | 6.0 | 49 (92) | 3 (5.6) | 0.5 ±0.4 | 1 (1.8) | 0.5 ± 0.0 |
|  | 7.1 | 51 (96) | 2 (3.7) | 0.2 ±0.1 | 0 (0) | NA |

### Clinical Interpretation, Positive and Negative Predictive Measures

While the DNN model has been formulated to output a single “days until labor” value, the MAE (difference between true and predicted) error used to measure model accuracy cannot be easily converted into clinically-meaningful measures for model positive and negative predictability. To aid clinical interpretation we introduced the concept of a prediction window *W*(*P*)_*T*_ derived from the error distribution of the model at time *T* with a given probability *P*. Table 3 gives an overview of model validation using a predictive labor window *W*(*P*)_*T*_ with *P* ∈ {0.7, 0.8, 0.9, 0.95} and *T* ∈ {7, 10}. The corresponding window sizes |*W*| for *T* = 7 where {3.7, 4.6, 6.0, 7.1}; at *T* = 10 where {6.2,7.4,9.2,10.4} days respective for each value in *P*. Given the limited sample size of our cohort we used all participants with a spontaneous birth for this analysis (N=54). We define true positive (TP) as the number of participants (N) whose labor correctly started within the window predicted by the model. False Positives (FP) are defined as those participants whose labor occurred after the model prediction window. For these individuals the model falsely predicts their labor to start in an earlier window as compared to true labor. We argue that this has a lesser impact on patient risk (they prepare earlier for labor to start if following model prediction window) when compared to False Negatives (FN). We defined false negatives as those participants who go into labor before the predicted labor window by the AE-LSTM models. These participants would potentially be unprepared for labor if following model predictions. For both FP and FN we also report by how much time (days) the model prediction missed the true labor. This is calculated as the difference between true labor and edge of the prediction window (right edge for FP, left edge for TP). This is indicated by “False Positive Window Days” and “False Negative Window Days” respectively in Table 3. We observe that our model can predict with a 79% TP, 18.8% FP and a small 1.8% FN rate 10 days prior to actual labor when given a window size of 7.4 days. Naturally increasing the window size, also increases our TP, and reduces both FP and FN, however a larger window size (ex: |W|=9.2, TP=96%) may not give pregnant mothers enough specificity to plan for labor. Similarly, we see a TP=79%, FP=15%, FN=5.6% when using a 4.6-day predictive window, 7 days prior to true labor.

## Discussion

The purpose of this study was to examine the feasibility of using continuously measured skin temperature to predict the onset of human parturition. We observed a common reduction in finger temperature of ∼ 0.5° C in the week prior to labor onset comparable to those occurring in other species (**Table 1**). In addition, we observed a reduction in the amplitude and stability of daily temperature rhythms, which appears to also occur in rodents^39^ and cows.^8^ We further related changes in the patterning of continuous body temperature to known hormonal changes preceding labor, suggesting that changes in thermoregulation reflect changes in hormonal state, as in other phases of female reproductive life.^11,12^

Notably, the present study demonstrates that applying deep learning techniques to continuous body temperature data enables accurate prediction of the day of labor onset. Our final model predicted labor onset 1 week prior to labor with an average signed error of <1 day, and a 79% certainty window of 4.6 days. As expected, we found that model error was greater in induced labors/planned Cesarean births, for which date of labor onset was *not* as related to the individual’s physiology but instead a product of complications, late gestational age, or a scheduled convenience. Indeed, it is likely that many induced labors would have begun naturally days after the induction was scheduled. In agreement with this, we observed that one prominent feature of approaching labor, lower body temperature, was not as low in induced labors, suggesting that induced pregnancies may be at a physiologically earlier state of development on average. We also observed that our predictions in induced labors tended to be late: induction occurred prior to when the model expected the mother to labor *physiologically.* However, the need for induction of labor in many cases indicates that pregnancy was not progressing along a healthy trajectory. In line with this, we also observed both more error *variability,* and reduced circadian power, which is typically associated with worse health across a variety of measures.^65–67^ Finally, we observed that model error tracked with gestational age in induced labors, as opposed to error remaining consistent across gestation prior to spontaneous labors. It is possible this indicates that, in absence of normally progressing changes in temperature, gestational age is the next most salient “feature” to attend to. Together, we propose that the combination of the dense physiological time series with the novel modelling approach enabled the application of an animal husbandry technique to the complex world of human pregnancy.

We recently demonstrated that *daily metrics* of average autonomic activity, physical activity, and sleep (including a single derived metric of body temperature) are useful in anticipating if labor onset will occur prior to or following a traditionally derived due date.^55^ Among these outputs, temperature provided the greatest contribution to the model. Although we were not able to find recent other recent studies of human temperature in labor prediction,^52^ a research group recently^68^ demonstrated decreases in physical activity with advancing gestational age, and an 18-participant cohort study demonstrated decreases in HR and increases in HRV in the third trimester.^69^ Although more human studies are needed, these results build on a wealth of animal literature demonstrating unique decreases in temperature prior to parturition, and numerous efforts to identify features indicative of imminent labor, ranging from simple thresholds^43^ to machine learning approaches similar to the present study.^70^ These low error rates, if confirmed in a larger clinical trial, would constitute a substantial improvement in pregnancy monitoring, and greatly improve families’ and clinicians’ ability to plan for the impending birth. To our knowledge, this is the first attempt to utilize continuous temperature alongside clinical and hormonal data for the purposes of anticipating labor onset in human pregnancy.

### Variability in Gestational Length

Interestingly, as gestation length varies fairly widely in humans, it is also unknown how far in advance the maternal and fetal bodies program labor and, accordingly, how far in advance it is possible to anticipate labor using any physiological signal. Some non-modifiable factors impacting gestational length include the shorter gestation of female fetuses^71^ and longer pregnancy in older or nulliparous women.^50^ There is also evidence that individuals who tend to have postdate, or preterm pregnancies will have a recurrence in subsequent pregnancies — suggesting a genetic effect on length of gestation.^71^ Indeed, each individual pregnancy is also an adaptive process. Labor onset timing may be influenced by local factors including infection exposure,^72^ stress,^73^ activity,^74^ maternal characteristics (body habitus^75–77^ or auto-immune diseases^78^) and timing of light exposure.^79^ The fetal CNS and adrenal maturation likely play a significant role, and are influenced in-part through placental hormonal production (CRH, estriol, progesterone), sterile inflammation,^80,81^ pro-inflammatory cytokines and prostaglandins (reviewed in^32,82^). Each of these changes may affect maternal physiological adaptation and time series. Complexity in the portrait of human labor physiology is likely due to overlapping mechanisms that may manifest different patterns person-to-person. As a result, any single feature of body temperature (or other vital sign) is likely insufficient for predicting human birth. Regarding temperature time series, which attributes of signal change are relevant to impending labor, and which are random in a population is something deep learning models are designed to determine.

### There are currently no reliable methods for clinical prediction of human labor

The current clinical estimate of delivery, the EDD, has an average error rate measured in weeks rather than days.^50^ The duration of ‘term’ pregnancy spans 5 weeks - from 37 to 42 weeks. There are no tools to help guide clinicians or pregnant individuals to predict if a pregnancy is likely to begin on the earlier or later side of this range. Preterm birth prediction remains elusive as well, though new tools are emerging using ML driven methods multi-omic data.^83,84^ A biomarker currently in use is the measurement of fetal fibronectin^85^ from a swab of the posterior fornix with a speculum examination. This is sometimes used in conjunction with ultrasound examination of cervical length.^86^ However, fetal fibronectin has a poor positive predictive value, and is only indicated in the presence of symptoms or risk factors—in addition to requiring a clinical encounter and discomfort with an internal examination which may be a barrier to timely assessment.

Presently, individuals are told to report symptoms of labor itself, which requires distinguishing vague symptoms of discomfort from true labor and yields high false-positive responses.^87,88^ Unfortunately, overt advanced symptoms of labor can occur without warning. This does not afford adequate time to intervene in the setting of preterm labor, may lead to unplanned home birth, or prompt recommendations for earlier labor induction when the uncertainty of waiting for labor is too high (e.g., living far from a hospital, if other obstetric complications are emerging). An accurate predictor a week or more in advance would allow clinicians and mothers to make care plans for the safest possible birth outcomes before labor starts.

### Considerations for future model development

The analyses presented here are a first attempt to combine machine learning and continuous skin temperature to anticipate labor in humans, alongside hormonal time series to validate the physiological basis of our findings. However, many challenges remain before such a model would be performant in a different, real-world cohort including pregnancies with more co-morbidities (e.g., gestational diabetes), those at risk for preterm birth, and a wider sociodemographic sample. Further validation will be necessary to determine how hormonal and temperature patterns differ depending on health risk-factors, and separate models may be needed to accurately make predictions in these mothers. Continuous data-based approaches are also hindered by the requirement to wear a device, such as a ring or bracelet, continuously. Large gaps in data will impact accuracy, and therefore limit the approach to those individuals willing and able to obtain, charge, and consistently wear a smart device. Future research is needed to determine the tolerance of this modeling approach to data gaps or data interpolation of more than a few hours.

## Conclusions

Continuous body temperature can be applied to anticipate labor onset with greater accuracy than the clinical standard. An AE-LSTM approach can extract relevant features of body temperature for accurate labor onset prediction from data across the third trimester. Features of temperature patterning, including temperature level and biological rhythms are correlated with changes in sex steroids over the final week of gestation. Future study of a larger population, including in high-risk pregnancies, will determine the broad clinical applicability of this approach.

## Data Availability

Data and Code Availability: The datasets used during the current study are available from the corresponding author on reasonable request. All code used to generate the findings here is available at - https://github.com/timebeforedelivery/laborprediction.git

## Acknowledgements

We acknowledge the contributions of time and data provided by the pregnant participants enrolled in this study, the research assistance from Sage Fannuci-Funes, CNM, DNP; and Gunjal Parekh for early data analysis. We would also like to thank Precision Analytical for working with us and our participants to promptly analyze hormone samples.

## Author Contributions

Study conception and design (EE, AG, SA, CB). Data analysis (CB, AG, SA, EE). Manuscript preparation (AG, EE, SA, CB). All authors read and approved the final manuscript.

## Data and Code Availability

The datasets used during the current study are available from the corresponding author on reasonable request. All code used to generate the findings here is available at - https://github.com/timebeforedelivery/laborprediction.git

## Supplemental Figures

**Supplemental Figure 1.**
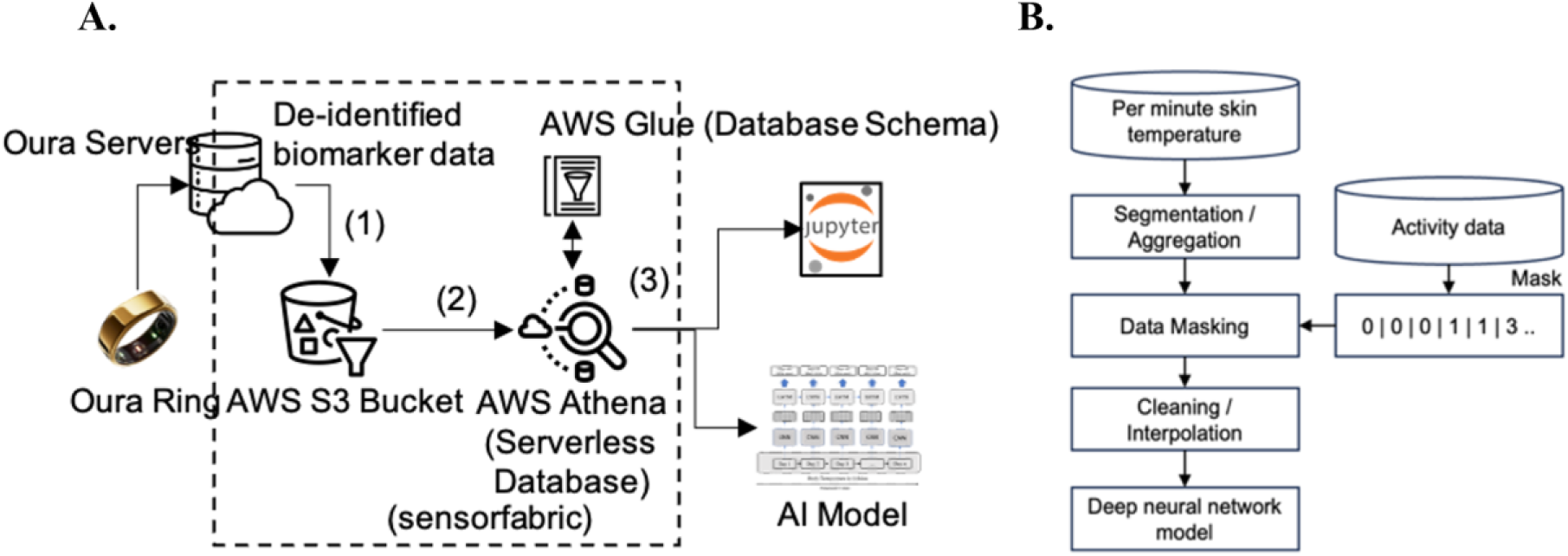
Data Preprocessing and Cleaning. Pre-Processing and Cleaning: data ingestion (A) and pre-processing pipelines (B). A): De-identified biomarker data from Ouraring, including high fidelity temperature and IBI are ingested into a campus secure Amazon Web Services (AWS) S3 bucket indicated by (1). Data is then parsed to generate structured schema, table meta-data in AWS glue, and participant partitions (to accelerate querying of per minute temperature data). These are then fed into a serverless querying solution provided by AWS Athena as shown by (2). B) We pass the raw minute temperatures from the device through pre-processing steps. First, we average the raw values over a 5-minute window, segment it into 24-hour periods starting at 10am each day, which allows the neural network model to learn from daily patterns (both day and night variation). Next, we remove data collected by the ring during non-wear time by using the 5 min activity labels provided by the ring, indicating wear/non-wear. Finally, we employed linear interpolation to account for missing and non-wear daily data. The final output is fed to a DNN model.

**Supplemental Figure 2.**
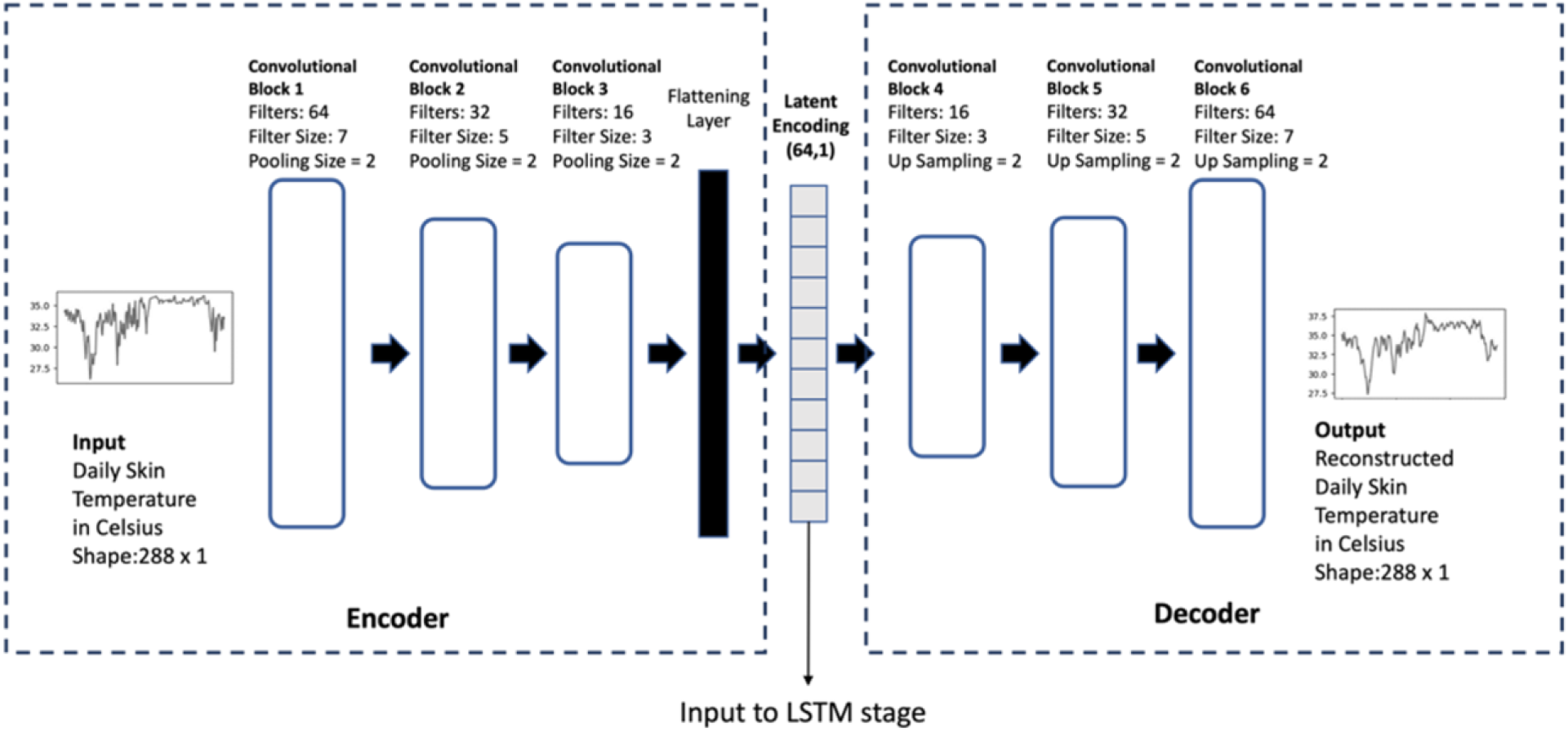
Autoencoder Structure. Auto encoders are divided into 2 parts – encoder and decoder. The encoder is responsible for converting values from feature space to latent space, while the decoder is responsible for converting them back to the feature space. AE train in an unsupervised setting where the object loss function MAE, measures the loss of reconstructing the original signal from the latent representation. In the encoder part of the AE, Input data ***T_k_*** of size 288 is fed into a series of three convolutional blocks. Each convolutional block comprises of a 1-D convolutional layer coupled with a max-pooling layer that enables reduction in data dimensionality. Output from the final convolutional layer is flattened and fed into a dense fully connected layer to produce the encoded representation. The decoder is a mirror image of the encoder. In the case of the decoder, the max-pooling layer is replaced by an up-sampling layer that gradually increases the dimensionality back to the original feature space.

**Supplemental Figure 3.**
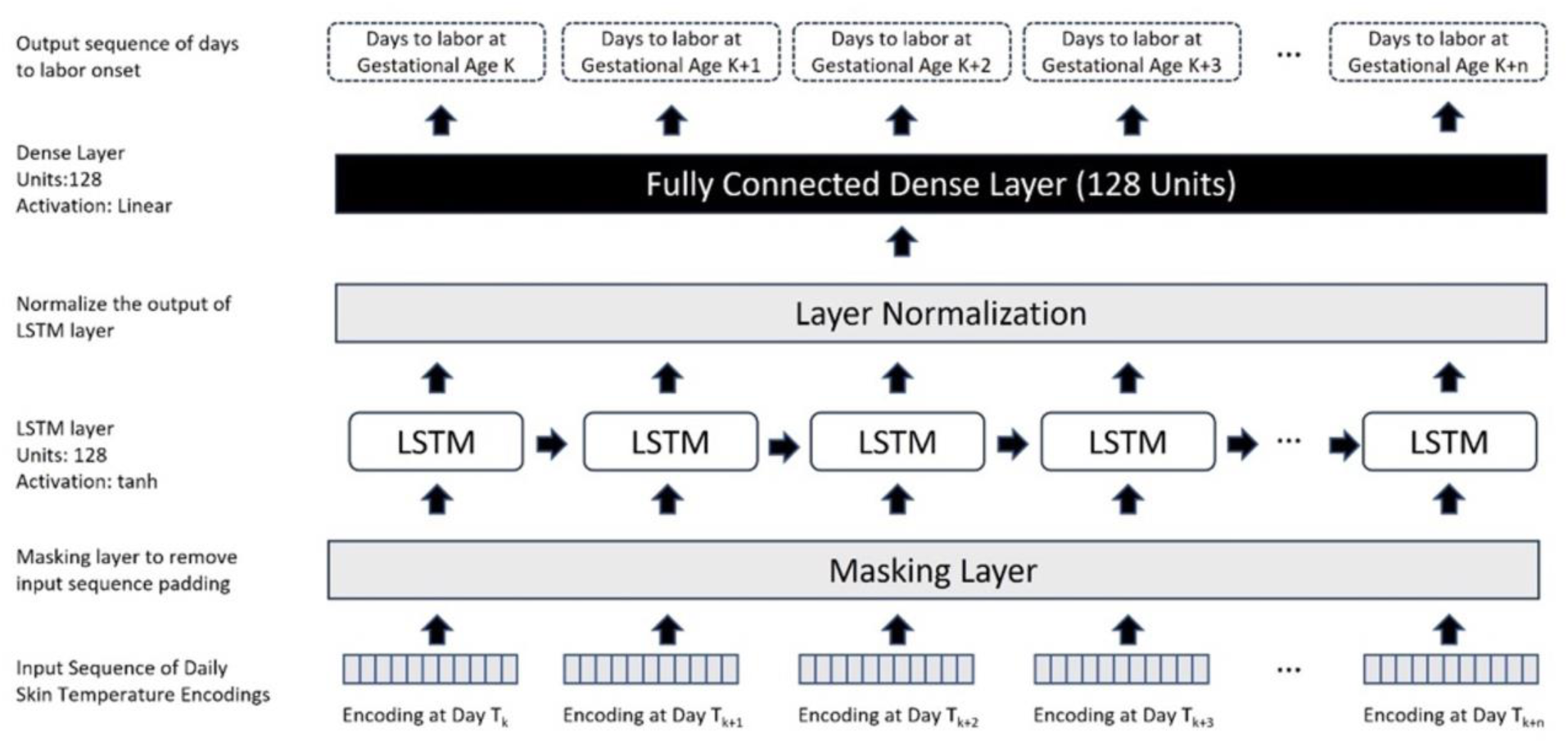
Long Short-Term Memory Model Structure. LSTM Stage (establishing the sequential relation): The LSTM model takes a sequence of 64-dimensional encoded vectors that represent daily skin temperature as input, and outputs days till labor relative to the current gestational age. We use zero-padding to conform the input sequences to a uniform length and the masking layer excludes zero values during analysis. The output of the masking layer is fed into an LSTM layer that is recurrent in nature. The LSTM layer has 128 units, and we use tanh as the activation function. We use layer normalization^102^ to normalize each output of the LSTM layer. Layer normalization reduces the dependency on batches, improves model performance, and is best suited for sequence-to-sequence models. Finally, the layer normalized output of the LSTM layer is fed into a dense layer with 128 units and linear activation function to output the days remaining to labor relative to current gestational age.

**Supplemental Figure 4.**
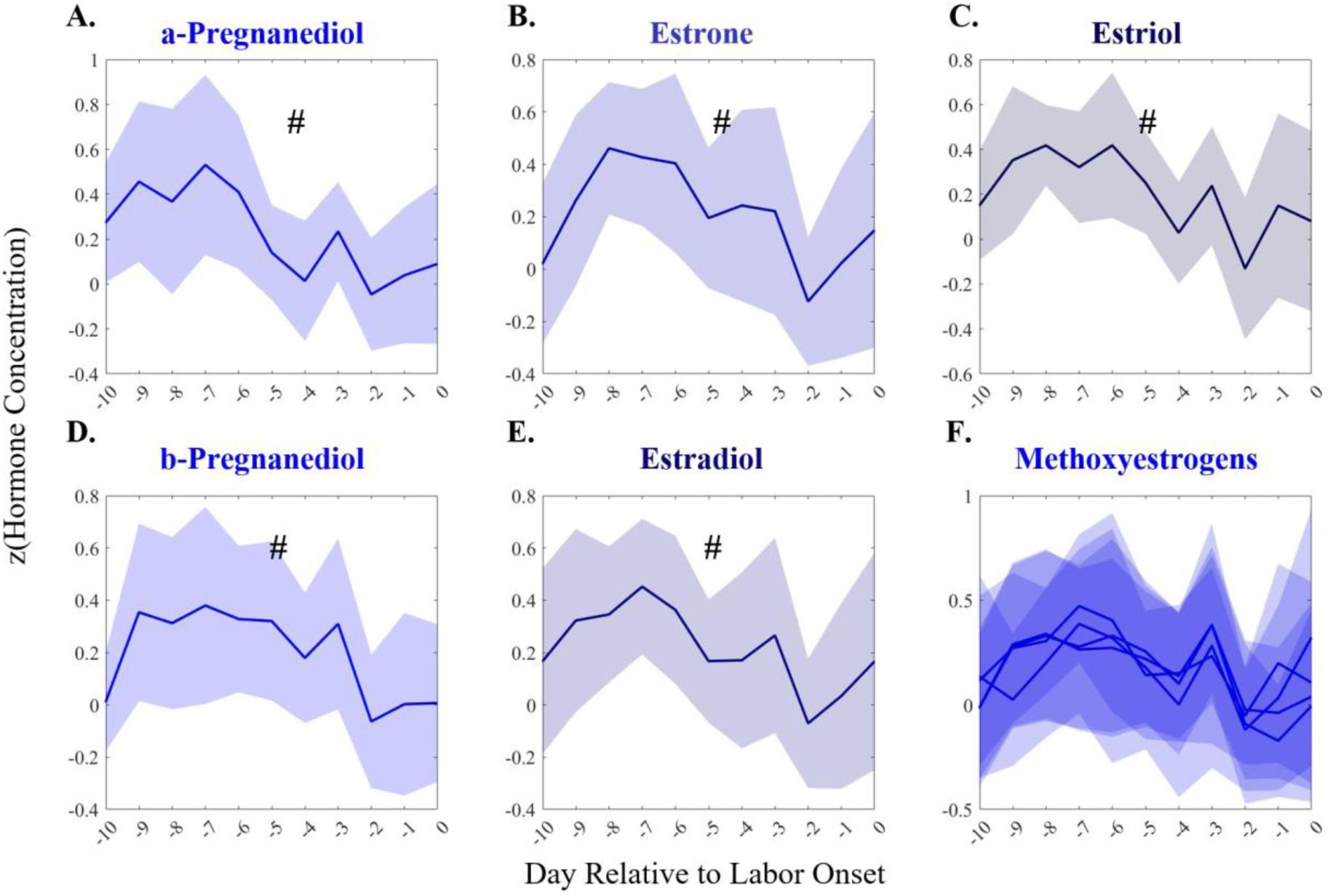
Spontaneous Labors Progesterone and Estrogen Metabolites Decrease in the 10 Days Prior to Labor Onset. Group means ± SEM of normalized hormone concentrations in the 10 days leading up to spontaneous labor onset (n=18). Estrogens, as well as α- and β-pregnanediol decreased across the 10 days prior to labor onset (# = p<0.05 in Mann-Kendall trend over time), a trend which disappeared or even reversed in the 2 days prior to labor onset (n.s.).

